# Shared genetic etiology between cortical brain morphology and tobacco, alcohol, and cannabis use

**DOI:** 10.1101/2021.03.28.21254282

**Authors:** Jill A. Rabinowitz, Adrian I. Campos, Jue-Sheng Ong, Luis M. García-Marín, Sarael Alcauter, Brittany L. Mitchell, Katrina S. Grasby, Gabriel Cuéllar-Partida, Nathan A. Gillespie, Andrew S. Huhn, Nicholas G. Martin, Paul M. Thompson, Sarah E. Medland, Brion S. Maher, Miguel E. Rentería

## Abstract

Genome-wide association studies (GWAS) have independently identified hundreds of genomic regions associated with brain morphology and substance use. However, the genetic overlap between brain structure and substance use has not been characterized. Here we leverage GWAS summary data of 71 brain imaging measures and alcohol, tobacco, and cannabis use to investigate their genetic overlap using LD score regression. We also used genomic structural equation modeling to model a ‘substance use common genetic factor’ and examined its genetic overlap with brain structure. After accounting for multiple testing, we identified eight significant negative genetic correlations, including between alcoholic drinks per week and average cortical thickness and intracranial volume with the age of smoking initiation; and five positive genetic correlations, including between insula surface area and lifetime cannabis use, and between the common factor with pericalcarine surface area. Our findings highlight a shared genetic etiology between variation in cortical brain morphology and substance use.

## INTRODUCTION

Heavy use of alcohol, tobacco, or cannabis is associated with serious negative consequences, including increased risk for unemployment,^1^ psychiatric comorbidity,^2,3^ substance use disorders,^4^ morbidity, and mortality.^5^ Over the last few decades, non-invasive brain imaging techniques have significantly bolstered our understanding of brain structure and function and their relationship with substance use. In parallel, lower genotyping costs, advancements in statistical genetics methods, and the availability of larger samples have enabled the investigation of genetic influences on both brain structure^6-8^ and substance use.^9,10^ Despite significant developments in neuroimaging and molecular genetics technologies, few studies have examined how the genetic architectures of alcohol, tobacco, and cannabis use are associated with neuroanatomical measures.

A substantial body of work has linked brain structural variation to substance use and abuse at the phenotypic level. For instance, cannabis use has been associated with reduced cortical thickness and surface area of the right entorhinal cortex,^11^ and an analysis of subcortical surface morphology identified localized differences in surface area and radial distance of the hippocampus, thalamus, putamen, and amygdala, among persons with alcohol dependence.^12^ The same study found surface area differences in the bilateral hippocampus, right nucleus accumbens, thalamus, and putamen of persons who use users.^12^ In another study, Gillespie et al.^13^ found an association between smaller thalamus volume and nicotine use in middle-aged males and no significant associations between subcortical volumes and cannabis use.^13^ Another study reported that higher levels of alcohol use were associated with thinner medial and dorsolateral frontal and parieto-occipital cortical regions, in addition to larger left ventricle volume.^14^ In two of the largest meta-analyses on substance use disorders and neurological structures to date, individuals with alcohol use disorder had lower cortical thicknesses in several brain regions (e.g., insula, precuneus)^15^ and smaller cortical volumes of the thalamus, putamen, hippocampus, amygdala, and accumbens.^16^ Taken together, although these studies vary widely in terms of their image acquisition methods and selection of regions, they highlight that alcohol, tobacco, and cannabis use are related to differences in brain morphology. However, it is unclear to which degree those relationships are underpinned by genetic factors.

The heritable nature of brain structure has been well-documented,^17-20^ with genetic influences explaining close to 70% of the variance in global, subcortical, and ventricular volumes; and 45% of the variance in frontal, parietal, occipital, and temporal lobe thickness.^20^ The genetic variation observed in brain structure and function is driven mainly by polygenic influences (i.e., multiple genetic loci of small individual effect size).^21-23^ Several studies to date have attempted to identify (a) common genetic variants underpinning brain structure, and (b) genetic variants associated with psychiatric conditions and their relationship with variability in brain morphology. In a recent genome-wide association meta-analysis, common genetic variants accounted for 34% and 26% of the variance in total cortical surface area and average cortical thickness, respectively.^6^ Also, previous studies have found either no or small negative genetic correlations between psychiatric phenotypes (e.g., schizophrenia, bipolar disorder) and intracranial volume.^24^ To our knowledge, only one study has investigated genetic correlations between substance use phenotypes and brain structural properties. This study found that cigarette smoking frequency showed a very small genetic correlation with total cortical surface area across the brain.^6^

Investigating whether the genetic architectures underpinning tobacco, alcohol, and cannabis use are related to brain morphology features could reveal brain-based pathways that influence gene-substance use relationships. With this in mind, the primary goal of the present study was to investigate potential shared genetic etiology between substance use and brain morphology. To accomplish that, we leverage the availability of recent genome-wide association studies (GWAS) summary statistics from large meta-analyses conducted by the Enhancing Neuro-Imaging Genetics through Meta-Analysis (ENIGMA) consortium,^7,25,26^ the GWAS & Sequencing Consortium of Alcohol and Nicotine use (GSCAN)^10^ and the International Cannabis Consortium^27^ to estimate pairwise genetic correlations between brain morphology measures (regional cortical surface area and thickness for 34 regions of interest and intracranial volume, total cortical surface area and mean thickness); and tobacco use measurements (age of smoking initiation, ever being a regular smoker, smoking heaviness and continuation vs. cessation), alcohol use (number of drinks per week), and lifetime cannabis use. Furthermore, consistent with the common liability hypothesis,^28^ genetic variants implicated in multiple substance use behaviors may be associated with specific or global neuroanatomic differences, an empirical question that remains unanswered. To answer that question, we used genomic structural equation modeling (gSEM)^29^ to model a common genetic factor across substance use phenotypes (i.e., tobacco, alcohol, and cannabis) and evaluate its genetic correlation with cortical morphology measures.

## METHODS

### Datasets

#### Brain imaging measures

GWAS summary statistics for 71 neuroimaging measures were obtained through direct application to the Enhancing NeuroImaging Genetics through Meta-Analysis (ENIGMA) Consortium.^25,26^ We specifically used summary statistics from the principal meta-analyses in European ancestry individuals for 68 bilateral cortical measures (thickness and surface area) and mean cortical thickness and total cortical surface area^6^ and a large GWAS conducted on intracranial volume.^30^ These measurements were based on brain MRI scans and genome-wide genotype data from the largest genetic studies of brain structure (Table 1). Participants in all cohorts from those studies gave written informed consent, and sites involved obtained approval from local research ethics committees or Institutional Review Boards. In the original analysis, GWAS summary statistics from each of the 50 sites had been combined using a fixed-effect inverse variance weighted meta-analysis in METAL.^31^

**Table 1.** Source of GWAS summary statistics datasets for phenotypes analyzed in this study.

| Study | Phenotypes | N participants | N cohorts |
| --- | --- | --- | --- |
| Hibar et al, 2017 | Intracranial volume | 33,536 | 65 |
| Grasby et al, 2020 | Mean cortical thickness<br>Total cortical surface area<br>Surface area and thickness for 34 ROIs | 33,992 | 50 |
| Liu et al, 2019<sup>a</sup> | Drinks per week<br>Ever regularly smoked<br>Smoking heaviness<br>Smoking cessation<br>Age at smoking initiation | ~1.2 million | 29 |
| Pasman et al, 2018<sup>a</sup> | Lifetime cannabis use | 184,765 | 18 |

**Note.** Summary statistics from the studies above only included European ancestry cohorts.
<sup>a</sup>The GWAS summary results also included the 23andMe cohort.

**Table 2.** Significant genetic correlations after multiple testing correction between brain imaging and substance use phenotypes.

| Neuroimaging Traits | Alcohol (Drinks Per Week) |  |  | Ever Regularly Smoked |  |  | Age of Smoking Initiation |  |  | Lifetime Cannabis Use |  |  | Common Substance Use Factor |  |  |
| --- | --- | --- | --- | --- | --- | --- | --- | --- | --- | --- | --- | --- | --- | --- | --- |
|  | rg | se | p | rg | se | p | rg | se | p | rg | se | p | rg | se | p |
| Intracranial volume | -- | -- | -- | -0.09 | 0.03 | 0.038 | -0.21 | 0.06 | 0.027 | -- | -- | -- | - | -- | -- |
| Postcentral gyrus surface area | 0.17 | 0.04 | $5.2 \times 10^{-4}$ | -- | -- | -- | -- | -- | -- | -- | -- | -- | -- | -- | -- |
| Total cortical surface area | 0.12 | 0.04 | 0.023 | -- | -- | -- | -- | -- | -- | -- | -- | -- | -- | -- | -- |
| Inferior temporal gyrus surface area | -- | -- | -- | -0.13 | 0.03 | $1.01 \times 10^{-3}$ | -- | -- | -- | -- | -- | -- | -0.13 | 0.03 | $1.6 \times 10^{-3}$ |
| Precuneus surface area | -- | -- | -- | 0.10 | 0.03 | 0.002 | -- | -- | -- | -- | -- | -- | 0.10 | 0.03 | 0.012 |
| Insula surface area | -- | -- | -- | -- | -- | -- | -- | -- | -- | 0.17 | 0.05 | .009 |  |  |  |
| Pericalcarine surface area | -- | -- | -- | -- | -- | -- | -- | -- | -- | -- | -- | -- | -0.09 | 0.03 | 0.012 |
| Average cortical thickness | -0.10 | 0.04 | 0.047 | -0.09 | 0.03 | 0.020 | -- | -- | -- | -- | -- | -- | -0.09 | 0.03 | 0.023 |

#### Tobacco and alcohol use measures

GWAS summary statistics for alcohol and tobacco use phenotypes were obtained from the repository of their corresponding publication,^10^ which included over 1 million individuals. Several GWAS were conducted, including (a) ever regularly smoked; (b) age of smoking initiation; (c) smoking heaviness (i.e., packs per day); (d) smoking cessation (i.e., current smoker versus former smoker); and (e) alcohol frequency (i.e., number of drinks per week). Specifically, GWAS were conducted among European ancestry individuals and included samples from both the 23andMe and GSCAN cohorts. Summary statistics for the 23andMe cohort were obtained via an application and signing of a data transfer agreement between 23andMe, Inc. and the QIMR Berghofer Medical Research Institute, where the genetic analyses were conducted.

#### Lifetime cannabis use

The GWAS summary statistics for lifetime cannabis use were retrieved from a meta-analysis (*N* = 184,765), which included European ancestry individuals from different cohorts such as The International Cannabis Consortium, UK Biobank, and 23andMe.^9^ Summary statistics excluding the 23andMe cohort were obtained from the International Cannabis Consortium’s online repository (https://www.ru.nl/bsi/research/group-pages/substance-use-addiction-food-saf/vm-saf/genetics/international-cannabis-consortium-icc/). Summary statistics for the 23andMe cohort were obtained via application and signing of a data transfer agreement between 23andMe, Inc. and the QIMR Berghofer Medical Research Institute, where the genetic analyses were conducted. Across all samples, participants reported whether they had ever used cannabis or marijuana (e.g., weed, dope, draw) in their lifetime. The 23andMe summary statistics datasets were meta-analyzed with the corresponding summary statistics from GSCAN and The International Cannabis Consortium’s datasets. We used an inverse variance weighted meta-analysis implemented in METALv2011-03-25.^31^

### Statistical Analyses

#### Linkage Disequilibrium Score Regression (LDSC)

Linkage disequilibrium score (LDSC) regression^32^ was used to assess pairwise genetic correlations between substance use (i.e., tobacco, alcohol, and cannabis use) and intracranial volume, as well as global and regional cortical surface area and thickness. LDSC leverages the expected relationship between the amount of LD that a variant tags and its association with a trait to model heritability and co-heritability using only the distribution of variant effect sizes. LDSC regression can then assess whether inflation in GWAS test statistics is due to polygenicity or confounding biases such as cryptic relatedness or population stratification. Likewise, bivariate LDSC regression can be used to distinguish between true genetic correlations between traits and inflation due to sample overlap. For this study, each dataset was filtered to only include markers overlapping with HapMap Project Phase 3 SNPs (N_overlap_□=□1,217,312), as these tend to be well-imputed across datasets, and alleles will match those listed in the data used to estimate the LD score. We used pre-computed LD scores for European populations, as provided on the LDSC website (https://github.com/bulik/ldsc). Standard errors were estimated using a block jackknife procedure and used to calculate p-values.

#### Genomic Structural Equation Modeling (GSEM)

To gain insights into the genetic etiology of substance use in general, we performed a common factor GWAS using genomic structural equation modeling (genomicSEM)^29^ implemented in R. This approach leverages the genetic variance-covariance matrix between the traits under study estimated through LDSC regression. Then, structural equation models can be used to partition the covariance structure and estimate latent factors. In this case, we desired to study the common genetic etiology of substance use phenotypes and thus specified a common factor model. The effect of a genetic variant on the common factor can be estimated by including the SNPs covariance with the traits studied in the model. Repeating this procedure for all genetic variants yields a GWAS of the common factor. Genetic correlations between this common factor GWAS and the neuroimaging traits of interest were performed using bivariate LDSC regression.

#### Brain Plots

The cortical thickness and surface area results are presented by mapping the z-score for the genetic correlation between a given trait and a brain region onto a brain triangular surface plot (see Figure 1). These plots were generated using python v.3.5 and the modules matplotlib, *numpy, plotly*, pandas, and *scipy*. All of the z-scores are shown without any filtering. However, statistically significant results are displayed on tables.

**Figure 1.**
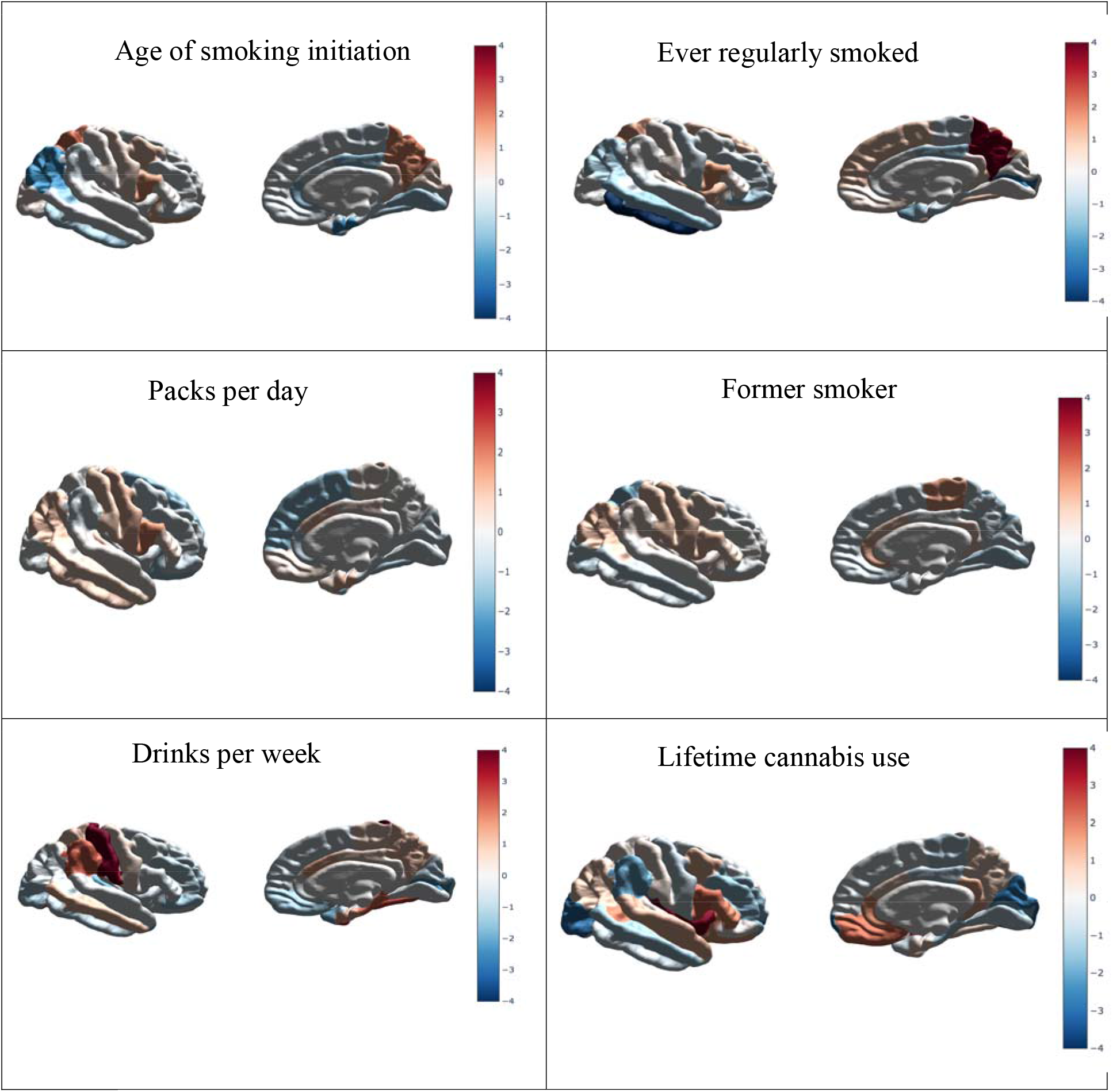
Standardized effect sizes (z-scores) reflecting the relationship between genetic risk for the substance use phenotypes and surface area. Positive effects highlighted in red denote increases in surface area and negative effects highlighted in blue denote reductions in surface area for coloured regions.

#### Multiple testing and significance threshold

We applied Benjamini-Hochberg’s False Discovery Rate (FDR < 5%) to account for multiple testing (i.e., the number of cortical neuroimaging traits) within each substance use phenotype.^33^ Genetic correlations that were nominally significant (i.e., *p* <.05) but did not survive multiple testing corrections are reported in the supplementary materials.

## RESULTS

### LD Score Regression Analyses

#### Alcohol

After correcting for multiple testing, significant positive genetic correlations were observed between alcohol use and total cortical surface area (*r*_g_ = 0.12; *p*-value = 0.023) and surface area of the postcentral gyrus’s regional area (*r*_g_ = 0.17; *p*-value = 4.3×10^-4^). A significant negative genetic correlation was observed between alcohol use and global average cortical thickness (*r*_g_ = −0.10, *p*-value = 0.047).

#### Smoking

Significant negative genetic correlations were observed between ever regularly smoked and (a) intracranial volume (*r*_g_ = −0.09, *p*-value = 0.038), (b) average cortical thickness (*r*_g_ = −0.09, *p*-value = 0.02), and (c) surface area of the inferior temporal lobe (*r*_g_ = −0.13, *p*-value = 8×10^-4^). A significant negative genetic correlation was observed between age of smoking initiation and intracranial volume (*r*_g_ = −0.21, *p*-value = 0.02). Last, a significant positive genetic correlation was observed between ever regularly smoked and precuneus surface area (*r*_g_ = 0.10, *p*-value = 0.002).

#### Cannabis

A significant positive genetic correlation was found between lifetime cannabis use and insula surface area (*r*_g_ = 0.17, *p*-value = 7.4×10^-3^). No other associations between cannabis use and neuroimaging traits remained significant after multiple testing correction.

#### Common Substance Use Factor

Negative genetic associations were observed involving the common substance use factor and the surface areas of the inferior temporal gyrus (*r*_g_ = −0.13, *p*-value = 0.002) and pericalcarine (*r*_g_ = − 0.09, *p*-value = 0.012), and cortical thickness (*r*_g_ = −0.09, *p*-value = 0.023). In addition, a positive genetic correlation was observed involving the common substance use factor and the precuneus surface area (*r*g = 0.10, *p*-value = 0.012).

## DISCUSSION

Although alcohol, tobacco, and cannabis use have previously been linked to brain structural differences,^11,12,14^ there is a dearth of work that has examined to which degree the genetic architecture of substance use overlaps with that of brain structure. The present study examined whether genetic liability for alcohol, tobacco, or cannabis use is associated with cortical brain morphology. Such work has the potential to shed light onto the molecular and neurobiological basis of substance use behavior and addiction.

In terms of the relationship between alcohol use and brain morphology, we found a genetic correlation between alcoholic drinks per week and a thinner average cortical thickness. These findings are consistent with previous work indicating that individuals with alcohol dependence display decreased cortical thickness compared to non-alcohol dependent individuals.^34^ There is some evidence that reduced cortical thickness is associated with characteristics such as poorer executive function, which may exacerbate the risk of drinking more frequently. We also found that a genetic correlation between drinks per week and larger cortical surface areas and surface areas of the postcentral gyrus, in line with previous work showing that alcohol abuse is associated with structural differences in the bilateral postcentral gyrus.^35^ Future work should investigate the precise mechanisms that account for the association between alcohol consumption and these brain phenotypes.

Regarding the relationship between smoking phenotypes and neuroanatomical traits, intracranial volume displayed negative genetic correlations with smoking initiation (i.e., ever being a regular smoker) and age at smoking initiation. Average cortical thickness and the surface of the inferior temporal gyrus displayed negative genetic correlations, and precuneus surface area had a positive genetic correlation with ever being a regular smoker. Previous observational studies have identified a relationship between phenotypic smoking and structural variation in the inferior temporal cortex (a region implicated in object and face recognition)^36^ and the precuneus (a region involved in motor imagery, directing attention, and processing abstract mental images).^37^ For instance, an increased number of years of smoking has been associated with smaller cortical volumes in the left middle temporal gyrus and right inferior temporal gyrus^38^ and cortical perfusion levels in the left precuneus.^39^ These findings suggest that the genetic association between brain morphology and tobacco smoking depends mainly on the age at which the behavior started or having a lifetime history of smoking regularly, rather than on its frequency or being a former smoker.

Lifetime cannabis use was negatively genetically correlated with insula surface area. Our results are in line with previous research that has linked phenotypic cannabis use to the insula. Individuals who have greater genetic liability for using cannabis may use cannabis more frequently, which has been associated with cortical alterations.^40,41^ For example, work by Chye et al.^12,41^ indicated that adolescents who used cannabis showed reduced thickness in the bilateral insula, a brain region that has been closely linked to addictive behaviors, including craving and drug-seeking, interoceptive processing, response to reward, and impulsive decision making.^42,43^ Other work has shown that individuals who initiate cannabis use had a smaller insula surface area in the right hemisphere.^44^

We also found that the common genetic factor substance displayed positive genetic correlation with precuneus surface area and negative genetic correlations with average cortical thickness and surface areas of the pericalcarine and the inferior temporal gyrus. These findings are in line with the genetic correlations observed for individual alcohol, tobacco and cannabis. However, the correlation with pericalcarine surface area was unique to the substance use common genetic factor. The pericalcarine cortex, which is involved in visual and spatial processing of information,^45^ has previously been linked to impulsivity and sensation-seeking ^45,46^ and substance use.^47^ A study reported that individuals who use more than one substance (i.e., alcohol and tobacco) had thicker left pericalcarine cortices relative to controls. However, this association did not remain significant when controlling for alcohol use.^47^

Limitations regarding the interpretation of the genetic correlations presented here must be acknowledged. These include the potential influence of pleiotropic effects. It is established that genetic variants associated with a given trait can also be related to other attributes as well.^22^ A notable potential mediator is that of cognitive ability and educational attainment. Cognitive ability has been robustly linked to increased total surface area and ICV.^48-50^ but has also been negatively associated with substance abuse phenotypes.^51-53^ Additionally, recent studies have shown that a genetic propensity for higher cognitive ability or educational attainment is linked to variation in regional cortical morphology, particularly in the frontal and temporal lobes.^50,54^ It is thus plausible that the associations we observe may, in part, be mediated by educational attainment – especially as many of the identified regions for both sets of traits are involved in cognitive processing, impulse control, and decision making. For example, one study found that the surface area and thickness of the prefrontal, insula, and medial temporal cortices were significant mediators of the relationship between polygenic scores for intelligence and general cognitive performance in two independent cohorts.^54^ While we cannot exclude the possibility that some of our observations are influenced by cognitive ability, several of our observations are in contrast to what would be expected if the relationship was entirely driven by cognitive ability. For example, we observed a positive genetic correlation between alcohol use and total cortical surface area. Future studies would benefit from examining the relationship among cortical morphology, substance use and cognitive ability together. An additional limitation of the study was that the causal nature of the relationship between substance use and brain morphology cannot be concluded. For example, differences in brain structure may predispose individuals to use substances and play a role in substance use reinforcement. Future work should consider employing methods to determine potential causal relations between drug use behaviors and brain structure.

Nevertheless, this study is the first to examine genetic correlations between tobacco, alcohol, and cannabis use with neuroimaging traits, illuminating the intricate relationship between the genetic architecture of substance use and brain structure. While these results suggest that genetic liabilities for substance use are associated with brain structure differences, there currently remains much to be understood about these relationships. With the increasing emphasis on precision medicine and personalized health initiatives, our work is a first step in helping to elucidate the complex relationship between genes, brain morphology, and behavior. In terms of the following steps, future research should consider examining genetics, brain structure, substance use and related behaviors, and environments over time to help determine the causal relationships among these variables. Such work has the potential to contribute to better understanding the mechanisms involved in the pathogenesis of substance use disorders, enable the detection of individuals at heightened risk for substance use problems, and aid in more precise diagnosis and treatment.

## Data Availability

Data are available upon request from ENIGMA and 23andMe. Data from ICC and GSCAN are publicly available.

http://enigma.ini.usc.edu/

https://research.23andme.com/dataset-access/

https://ibg.colorado.edu/mediawiki/index.php/GSCAN

https://www.ru.nl/bsi/research/group-pages/substance-use-addiction-food-saf/vm-saf/genetics/international-cannabis-consortium-icc/

## ACKNOWLEDGEMENTS

We thank the research participants and employees of 23andMe for their contribution to this study. MER’s work was supported by the Australian National Health & Medical Research Council and the Australian Research Council through an NHMRC-ARC Dementia Research Development Fellowship (GNT1002821).

## DATA AVAILABILITY

The data that support the findings of this study are available from the corresponding author upon reasonable request. To apply and access 23andMe summary statistics, please visit research.23andme.com/dataset-access/ for more information.

